# Assessing Plasticity in the Primary Sensory Cortex and Its Relation with Atypical Tactile Reactivity in Autism: A TMS-EEG Protocol

**DOI:** 10.1101/2024.05.24.24307857

**Authors:** Shohreh Kariminezhad, Reza Zomorrodi, Christoph Zrenner, Daniel M. Blumberger, Stephanie H. Ameis, Hsiang-Yuan Lin, Meng-Chuan Lai, Tarek K. Rajji, Yona Lunsky, Marcos Sanches, Pushpal Desarkar

## Abstract

**Background:** Atypical sensory reactivity is a cardinal presentation in autism. Within the tactile domain, atypical tactile reactivity (TR) is common, it emerges early, persists into adulthood, and impedes social interaction and daily functioning. Hence, atypical TR is a key target for biological intervention to improve outcomes. Brain mechanisms informing biological interventions for atypical TR remains elusive. We previously reported hyper-plasticity in the motor cortex in autistic adults and found that repetitive transcranial magnetic stimulation (rTMS), designed to strengthen inhibitory processes in the brain, reduced hyper-plasticity. Whether the primary sensory cortex (S1) is characterized by hyper-plasticity, which may underlie atypical TR in autism is unknown.

**Objectives:** We aim to test whether hyper-plasticity in the S1 underlies atypical TR in autism, and investigate if a single session of rTMS can safely reduce hyper-plasticity in S1 in autistic adults.

**Method:** Plasticity will be assessed in the left S1 with integrated paired associative stimulation and electroencephalography (PAS-EEG) paradigm in 32 autistic adults and 32 age-, sex-, and intelligence quotient-matched controls. Autistic participants will be further randomized (double-blind, 1:1) to receive a single-session of either sham or active 20 Hz bilateral rTMS over the S1 and the plasticity will be re-assessed over the left S1 on the same day.

**Conclusions:** Atypical TR has been identified as one of the top clinical research priorities that can influence outcome in autistic population. The study findings can be highly valuable to further elucidate the mechanism underlying atypical TR, which in turn can help with developing a mechanism-driven intervention.

## Introduction

Autism Spectrum Disorder (hereafter will be called autism) is one of the most common neurodevelopmental conditions, affecting approximately 1% of the world population (1). It is estimated that over 90% of autistic individuals exhibit atypical sensory reactivity (2). Atypical sensory reactivity in the form of hyper- or hypo-reactivity to external stimuli is a cardinal presentation in autism. In the sensory domain, atypical tactile reactivity (TR) is a common presentation that emerges early, persists into adulthood, and adversely affects social interaction and daily functioning, thus significantly contributing to the overall disability (3,4). An international commission on the future of care and clinical research in autism identified the sensory domain as one of the top clinical research priorities that can influence care and outcome in autism (5). We engaged autistic adults attending our specialized autism clinic and received consistent feedback that this is a high-priority area of significant unmet need. Behaviorally, both tactile hypo- and hyper-reactivity are believed to be on the same continuum reflecting the same underlying biological process where hypo-reactivity is a coping mechanism to deal with excessive stimulation (6). Neurophysiologic studies of tactile processing (4,6) and neuroimaging studies of excitatory and inhibitory metabolites in the primary sensory cortex (S1) in autism remain inconsistent and inconclusive (7,8); therefore, brain processes informing biological interventions for atypical TR remains elusive.

Converging evidence indicates that the neurobiology of autism is characterized by atypical plasticity. A key insight from well-characterized valproic acid animal models of autism is that an excessive long-term potentiation (LTP) plasticity or hyper-plasticity adversely affects behavior (9–11). Hyper-plasticity was found in the S1 of mice that displayed atypical TR (11). Whether the S1 is characterized by hyper-plasticity, which may underlie atypical TR in autistic humans, is unknown.

A more direct evidence of hyper-plasticity was consistently observed in human motor cortex using transcranial magnetic stimulation (TMS) (12–15) with one exception (16). Our group replicated this finding of hyper-plasticity in the motor cortex in autistic adults (15). As a foundation for intervention, we also collected pilot data using a repetitive transcranial magnetic stimulation (rTMS) protocol designed to strengthen inhibitory mechanisms, which reduced hyper-plasticity in autistic adults (15). In our previously published study (15), we conducted a randomized trial involving 29 autistic adults. The participants were allocated (1:1) to undergo either a single session of active or sham rTMS, with 6,000 pulses applied at 20Hz, targeting the motor cortex. In comparison to other widely-used inhibitory rTMS protocols, that are 1Hz and 10Hz, Dejesus et al. showed that 20Hz rTMS, when administered with 6,000 pulses, produced the maximum inhibitory effect (17). The results revealed a substantial effect size of active rTMS on Long-Term Potentiation (LTP), with LTP reduction observed on the day following rTMS. This decrease in hyper-plasticity aligns with the altered neuronal excitation/inhibition (E/I) model of autism (18). According to this model, the hyper-plasticity observed in autism is associated with an increased E/I ratio, and facilitating inhibition could contribute to the observed reduction. The rationale behind such approach has been previously published by our team (19).

Our primary goal is to test whether the S1 hyper-plasticity underlies atypical TR in autism. In this study, we will use the innovative combination of paired associative stimulation (PAS), a well-established TMS paradigm to study plasticity, and electroencephalography (EEG) or PAS-EEG, to assess plasticity (20). In PAS-EEG experiment, PAS induced LTP (i.e. PAS-LTP) is defined by potentiation of cortical evoked activity (CEA), which is measured by EEG. The reproducibility of the PAS-EEG method was reported in independent samples (21,22). We will also explore if a single session of ‘mechanism-driven’ rTMS (15) described above can reduce hyper-plasticity in the S1 and whether it is well-tolerated by autistic adults.

## Objectives

In this study, there are two primary objectives:

1. To assess and compare plasticity in the S1 between autistic adults and neurotypical controls.
2. To examine the relationship between TR and plasticity in the S1 within both study groups, i.e. autistic and neurotypical.

Additionally, we have two exploratory objectives:

1. To investigate if a single-session of active bilateral rTMS delivered to the S1 can reduce the S1 hyper-plasticity in autistic adults.
2. To assess tolerability of bilateral rTMS to the S1.

## Hypotheses

### Main hypotheses

Our main hypotheses, pertaining to objectives 1 and 2, are as follows:

Hypothesis 1: Compared to neurotypical controls, autistic adults will display hyper-plasticity (i.e. significantly greater LTP) in the S1.

Hypothesis 2: There will be a positive association between plasticity and TR, however, the association will only be significant for the autistic group reflecting hyper-plasticity underlying excessive TR.

### Exploratory hypotheses

Hypothesis 1: Autistic adults randomized to active bilateral rTMS will have less LTP in S1 following a single session of active rTMS than those randomized to single session of sham bilateral rTMS.

Hypothesis 2: Bilateral active rTMS to S1 will be well-tolerated.

## Method

### Design and Setting

The current study, comprising two phases (Fig 1), one of which is randomized and double-blind, will be conducted at CAMH’s Temerty Centre for Therapeutic Brain Intervention, Toronto, Canada, from April 2023 to April 2025. The authors assert that all procedures contributing to this work comply with the ethical standards of the relevant national and institutional committees on human experimentation and with the Helsinki Declaration of 1975, as revised in 2013. All procedures involving human subjects were reviewed and approved by the research ethics board at CAMH (135/2019).

**Fig 1.**
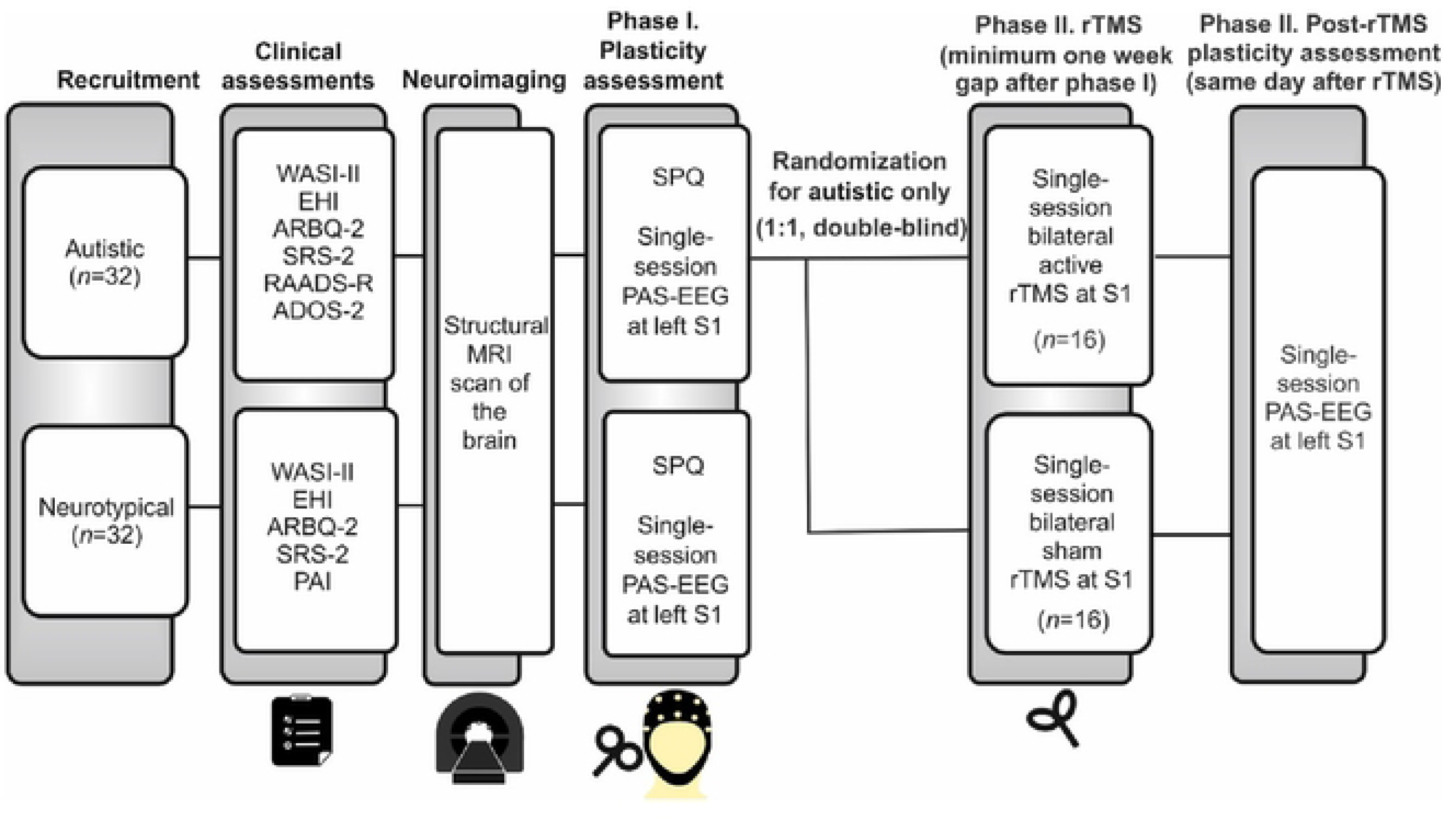
Schematic diagram of the study design. Initially, all participants (*N =* 64) will undergo a baseline clinical assessment and MRI brain scan. In their third visit, the participants will complete the SPQ questionnaire and the plasticity in the left S1 will be assessed using PAS-EEG. Only autistic participants will be further randomly assigned to either the active or sham group, in a double-blind manner, to receive bilateral rTMS over the S1, and the plasticity of the left S1 will be reassessed on the same day following rTMS. This session will be scheduled at least one week after the third visit. WASI-II, Wechsler Abbreviate Scale for intelligence - second edition; EHI, Edinberg Handedness Inventory; Edinberg Handedness Inventory (ARBQ-2); Social Responsiveness Scale-2 (SRS-2); Ritvo Autism Diagnostic Scale-Revised (RAADS-R); Autism Diagnostic Observation Schedule - second edition (ADOS-2); MRI: magnetic resonance imaging; SPQ: Sensory Perception Quotient; PAS: paired associative stimulation; EEG: electroencephalography; S1: primary sensory cortex; rTMS: repetitive transcranial magnetic stimulation.

All participants, both autistic and neurotypical, will undergo clinical assessments, which include diagnosis confirmation, handedness determination, IQ evaluation, and a TMS safety screen. Additionally, they will undergo a magnetic resonance imaging (MRI) scan of the brain, which will be used for MRI-guided neuronavigated TMS in subsequent sessions. On their next visit, all participants will complete the 92-item Sensory perception Quotient to assess TR (23). Then, plasticity will be assessed in the left S1 using PAS-EEG paradigm.

Upon completing the first phase, only autistic participants will be randomly assigned in a double-blind manner (1:1) to receive either a single-session of active or sham bilateral rTMS over the S1, followed by PAS-EEG plasticity assessment session, on the same day. Assignment will be determined using a permuted block randomization method generated by an independent biostatistician external to the project. The random allocation sequence will remain concealed throughout the experiment and data analysis, with only the investigators being unblinded upon finalization of the study.

The rTMS session will be scheduled with at least a one-week gap from the first plasticity assessment session to allow any potential carryover effects from the PAS to dissipate. This ensures that the subsequent rTMS treatment will not be influenced by the prior PAS intervention. The first participant was recruited on June 08, 2023. Fig 1 provides a schematic diagram illustrating the study design.

### Participants

This study aims to enroll a total of 64 participants, comprising 32 adults diagnosed with autism as per fifth edition of the Diagnostic and Statistical Manual of Mental Disorders (DSM-5), without intellectual disability (24). This diagnosis will be confirmed through clinical assessment and the Autism Diagnostic Observation Schedule-2 (ADOS-2) (25).

It is important to note that our study will encompass autistic participants who have stable co-occurring mental health conditions and medications (41). Research findings indicate that 70% of young autistic individuals have at least one co-occurring mental health condition (26–28), and almost half of them meet criteria for more than one (27,28). We believe that excluding individuals with co-occurring mental health conditions and medication use is impractical and would hinder the generalizability of our findings. In addition, given the normative sex differences in plasticity (29), we will recruit at least 11 females, reflecting a higher male:female (2:1) ratio compared to previous ratio of 2.5-3:1 (30).

Autistic participants will primarily be recruited from Adult Neurodevelopmental Services outpatient clinics at CAMH, and CAMH research registry.

For the control group, an additional 32 neurotypical participants will be recruited through the healthy control registry, alongside advertising at local universities and in newspapers. The recruitment process will involve a 1:1 matching strategy, ensuring that for each autistic adult, a corresponding neurotypical counterpart will be sought based on age, sex, and IQ.

All prospective participants, expressing their willingness to take part in this study, will be referred to our research analyst for a detailed discussion about the study’s objectives, protocols, potential risks, and their rights as research participants. Our research team will then assess the participant’s eligibility for inclusion in the study, with the inclusion and exclusion criteria detailed in Table 1. Each participant will be required to provide a written consent at the outset of the study.

**Table 1.**
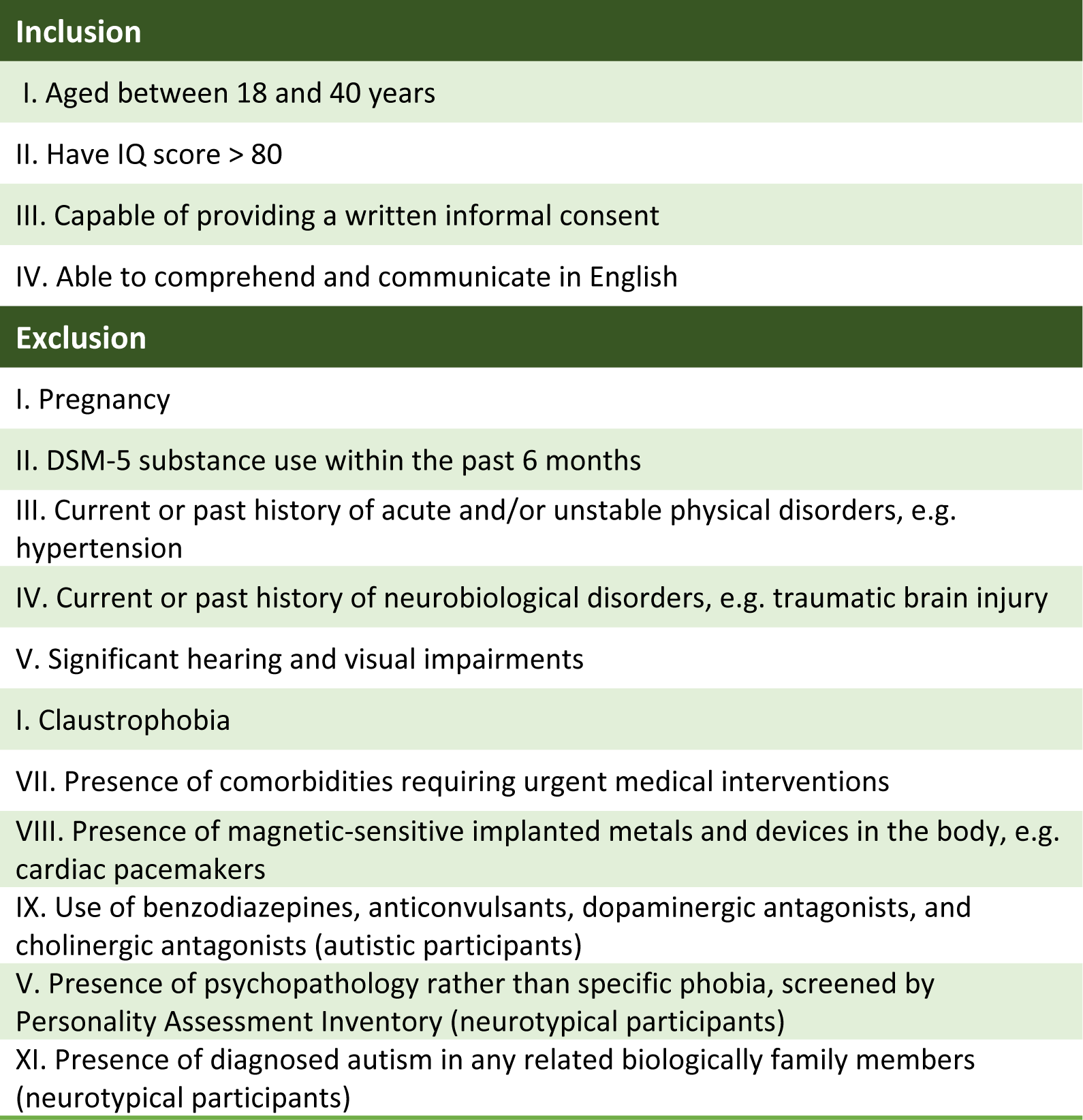
Inclusion and exclusion criteria.

### Measurement

#### Baseline Clinical Assessment

At the outset of the study, we will perform a series of baseline assessments. All participants who meet the inclusion/exclusion criteria will be administered shortened second edition of the Wechsler Abbreviate Scale for intelligence (WASI-II), Edinberg Handedness Inventory (EHI), Adult Repetitive Behavior Questionnaire-2 (ARBQ-2), the Social Responsiveness Scale-2 (SRS-2), and SPQ (31–34, 23). To screen for the presence of general psychopathology in control adults, neurotypical participants will also complete the Personality Assessment Inventory (PAI) (35). Furthermore, other than participating in ADOS-2, all autistic participants will complete the Ritvo Autism Diagnostic Scale-Revised (RAADS-R) (36).

#### Phase I. Assessment of Plasticity

##### Transcranial Magnetic Stimulation

TMS procedures will be performed using a Magstim 200 stimulator (Magstim Co., Whitland, UK) equipped with a 70-mm figure-of-eight coil. To enable neuronavigated TMS, structural T1-weighted magnetic resonance (MR) images will be obtained prior to the TMS session. These images will be acquired with a 3T MRI scanner (Discovery **MR750, GE** Healthcare, Chicago, IL, USA) for each participant at the Research Imaging Centre, CAMH. Subsequently, these MR images will be imported into BrainSight software (Rogue, Research Inc, Canada) for individual MRI-guided navigated TMS (nTMS).

The TMS stimulation session will begin with the identification of the cortical representation of the abductor pollicis brevis (APB) muscle, a process known as ‘hotspotting’. To locate the APB hotspot, we will position the intersection of the coil tangentially to the participants’ skull, optimizing it around the C3 electrode (the approximate location of the primary motor cortex for the hand). The coil’s handle will be oriented backward, forming a **45° angle** with the midline. Once the hotspot will be located, we will determine the resting motor threshold (RMT). The RMT will be identified as the lowest stimulus intensity required to induce MEPs with a peak-to-peak amplitude of at least 50 microvolts (µV) in the relaxed right APB muscle, in at least half of 10 consecutive trials. Subsequently, 120% of RMT will be used to determine the stimulus intensity of 1 mV (SI1_mV_), i.e. the intensity of TMS needed to produce MEPs with mean peak-to-peak amplitude of 1 mV over 20 TMS trials. TMS stimulation will be then applied to the left S1 site (MNI coordinates: X = −48 mm, Y = −21 mm, Z = 50 mm), with the intensity set at SI1_mV_ throughout the session. In total, 100 single pulses will be delivered. This paradigm will be administered both before and after the PAS intervention at different time points (0, 15, 30, and 60 minutes) (Fig 2.A).

**Fig 2.**
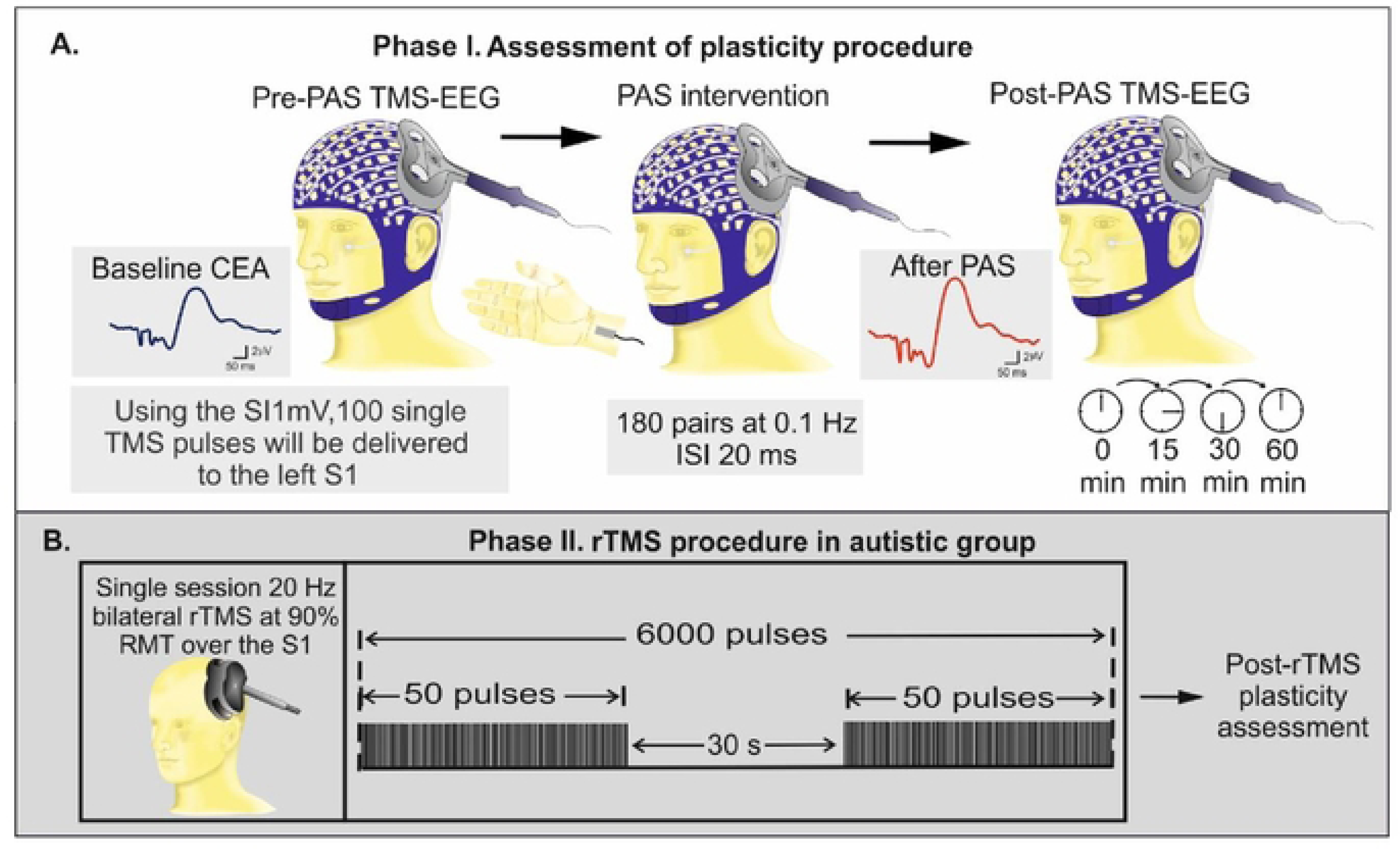
Schematic figure of plasticity assessment and intervention phases of the current study. (**A**). The plasticity assessment procedure consists of delivering 100 single TMS pulses over the left S1 using SI1 mV which will be applied before and after (0, 15, 30, and 60 min) PAS intervention. Throughout the PAS intervention, electrical stimulation of the right median nerve will precede TMS stimulation of the left S1 at an ISI of 20 ms. The plasticity will be then assessed using the ratio of the maximum CEA following PAS to the baseline CEA. CEA is quantified as the area under the curve of rectified TMS-EEG response within a time window of 25-275 ms from TMS stimulus onset. (**B**). Autistic participants will be further randomized to receive either a single session sham or active 20 Hz rTMS. The rTMS intervention will be delivered bilaterally over the S1 consisting of 120 trains of 50 pulses with an ITI of 30s (in total, 6000 pulses). The sequence of delivery will be randomized for each participant, that is left S1 followed by right S1 and vice versa, and the pulses will be applied at an intensity of 90% RMT. Following rTMS, on the same day, TMS-EEG plasticity assessment paradigm will be applied.

Participants will remain seated in a comfortable chair for the entire session, and to mask the auditory stimulation from TMS, white noise will be played through earplugs, with the volume adjusted at the limit of their comfort.

##### Electroencephalographic Recording and Preprocessing

EEG recordings will be continuously acquired concurrently with TMS using a TMS-compatible 64-channel amplifier (Compumedic Inc) while the electrode impedances will be kept below 5 kΩ. The EEG recorded data will be referenced against the electrode positioned posterior to the vertex electrode (Cz). The sampling rate of 20 kHz and a low-pass filter of 100 Hz will be selected, to minimize the artifacts introduced by TMS and avoid amplifier saturation (37).

Thereafter, the acquired EEG data will be preprocessed offline using customized scripts in MATLAB (MathWorks Inc., Natick, MA, USA) and EEGLAB open source MATLAB toolbox, in accordance with a defined cleaning pipeline. According to this pipeline, the EEG data will be (1) trimmed from −2 ms to 10 ms after TMS pulse; (2) decay artifact will be removed with ICA and then data will be down-sampled to 1 kHz; (3) segmented into epochs (−1000 ms to 1000 ms with respect to the TMS pulses); (4) baseline-corrected using the artifact-free pre-TMS interval (from −500 ms to −5 ms); (5) another round of ICA will be applied to remove residual artifacts such as EOG and muscle artifacts. In the last step, data will be filtered using a zero-phase shift band-pass filter with a frequency range of 1 to 100 Hz and60 re-referenced using an average reference for further analysis.

##### Paired Associative Stimulation Intervention

To induce plasticity in the S1, a PAS paradigm will be adopted. During this PAS paradigm, one hundred eighty electrical stimuli will be delivered at 0.1 Hz over the right median nerve. The intensity of these stimuli will be set to three times the participant’s perceptual threshold, which is the minimum intensity at which they can perceive the stimulation. To generate an LTP-like plasticity effect, the peripheral stimulation of the median nerve will be preceded the TMS stimulation of the contralateral S1 by 20 ms (Fig 2.A). The 20 ms interval will be employed to ensure that the timing of the peripheral nerve stimulation and TMS is aligned with the arrival of the peripheral sensory input in the S1 (38). Participants will be instructed to maintain their attention on their stimulated wrist and count the number of pulses they will receive throughout the protocol. This instruction is based on the recognition that attention can modulate the effects of PAS, as suggested by the study conducted by Stefan et al (39).

#### Phase II. Repetitive Transcranial magnetic Stimulation Procedure in Autistic Participants

Stimulation will be conducted with an X100 stimulator with a double-sided B65 A/P type fluid-cooled coil (MagVenture, Farum, Denmark Inc.) which is a combined active/sham coil with identical sides. Single-session 20 Hz rTMS will be delivered bilaterally over the S1 at 90% of RMT, with 120 trains of 50 pulses and an inter-train interval (ITI) of 30 s (Fig 2.B). In total, 6000 pulses will be administered to each autistic participant. The sequence of delivery will be randomized for each participant, i.e., left S1 followed by the right S1 or vice versa. Sham stimulation will be delivered using the sham side of the coil, which does not generate a magnetic field but mimics the sensation of magnetic stimulation and produces the same clicking sounds.

### Outcomes

#### Primary Outcome

The primary outcome measure of this study is the PAS-induced LTP in the S1. PAS-induced LTP will be assessed using CEA. CEA is quantified as the area under the curve of rectified TMS-EEG response within a time window of 25-275 ms from TMS stimulus onset (40). This chosen time window is designed to exclude any early artifacts and account for the subsiding effects of TMS. CEA will be averaged across 100 epochs from the electrodes of interest corresponding to the S1 region. The mean post-PAS CEA will be then divided by the pre-PAS CEA, reflecting the potentiation of CEA at each of post-PAS time points (0, 15, 30, and 60 minutes). Given that the timing of the post-PAS maximum potentiation could differ among participants, we will use the maximum CEA ratio for each participant for the statistical analysis (20).

To asses TR, the total score on the “touch” category of SPQ questionnaire will be used.

#### A *Priori* Power Calculation

A sample size of *N* = 64 participants, consisting of 32 autistic participants and 32 neurotypical controls, has been determined as adequate for hypothesis testing. This sample size provides 90% power. This calculation is based on the preliminary data, the observed LTP difference of 20.6 points, as evidenced in our PAS-EEG pilot study involving both autistic adults and control subjects. Moreover, the same sample size is deemed suitable for evaluating hypotheses 2, offering an 80% statistical power to detect a moderate effect size (Cohen’s f2 = 0.15). These power calculations were executed using G*Power.

Drawing upon our previously published findings (15), we envision a “substantial” impact of active rTMS in a group of 16 autistic participants when compared to a sham group also consisting of 16 autistic participants. It is important to note that, given the relatively small sample size, the application of a single session, and the exploratory nature of this outcome, our approach to presenting results and their interpretation will extend beyond statistical significance values. Instead, we will place significant emphasis on estimated effects and their precision, including the calculation of 95% confidence intervals. This comprehensive set of information, combined with data related to tolerability, will play a pivotal role in shaping the design of future clinical trials.

### Statistical Analysis

Hypothesis 1: To test this hypothesis, we will employ a linear regression model with CEA ratio (LTP) as dependent variable and study groups (autistic and neurotypical controls) as independent variable. Age, sex, hemisphere of PAS-EEG assessment, IQ and pre-PAS CEA (an index of individual baseline cortical excitability) will enter the model as ‘control’ independent variables. To ensure robustness, a model diagnostic will be conducted through analysis of residuals and Cook’s distance. Potential confounding effect of medication use among autistic adults on plasticity will be investigated in a post-hoc sensitivity analysis.

Hypothesis 2: We will use a multivariate analysis of variance to test this hypothesis where TR will be dependent outcome; S1 plasticity, study groups and their interaction will be the main predictors; and sex, age, hemisphere of PAS-EEG assessment and IQ will enter the model as controls. If the interaction between S1 plasticity and study group proves to be significant at confidence level 0.05, a further investigation of the nature of the association will be conducted separately within each group, by means of linear contrasts.

Exploratory Hypotheses: In order to study the effect of active vs. sham rTMS on LTP, we will use an analysis of covariance with CEA ratio (LTP) as dependent variable, groups (active and sham rTMS) as fixed factor, and pre-rTMS CEA ratio as covariate. We will count discontinuation of rTMS session and adverse events to assess tolerability.

## Discussion

Our study will be the first to assess plasticity in the S1 in autistic adults using the innovative PAS-EEG. Completion of our study objectives will fill in a critical void in our understanding, i.e., if successful, we will identify the S1 hyper-plasticity as one brain mechanism underlying atypical TR and associated behavioral characteristics of autism. This information will provide us a biological target for testing ‘mechanism-driven’ interventions for tactile sensory difficulties in autism. Further, S1 hyper-plasticity may serve as a potential biomarker for TR and core behavioral characteristics in autism.

In conclusion, this protocol describes the study rationale, objectives, hypotheses, design and proposed methodologies of identifying the S1 plasticity in autistic adults. This study may provide a new line of translational inquiry that has not been explored in autism before.

## Data Availability

No datasets were generated or analysed during the current study. All relevant data from this study will be made available upon study completion.

## Authors’ Contributions

S.K. participates in data acquisition, analyzing and interpreting the data, and wrote the first draft of the manuscript. P.D. is involved in study design and administration, data analysis and interpretation, and critically revised the manuscript. R.Z. is involved in study design, data analysis and interpretation, and provided critical revisions to the manuscript. M.S. is involved in data analysis. C.Z., DM.B., SH.A., HY.L., MC.L., TK. R., and Y.L., all contribute to data interpretation and critically reviewed the manuscript. The final manuscript was approved by all authors.

## Acknowledgment

This work is supported by the Innovation Fund from the Alternate Funding Plan of the Academic Health Sciences Centres of Ontario, CAMH Discovery Fund, CAMH Discovery Fund Postdoctoral Fellowship Award (SK), and the Academic Scholar Award from the Department of Psychiatry, University of Toronto. We extend our gratitude to the study participants for their contributions to this research. Additionally, we would like to thank Ashna Imran, and Sushmit Das, research analysts, for their assistance in data acquisition and management.

